# Effect of parents’ country of birth on the relationship between socioeconomic status and childhood overweight and obesity in Southwest Sydney, Australia: learnings from the Growing Healthy Kids population prevalence survey

**DOI:** 10.1101/2024.07.02.24309842

**Authors:** Vilas Kovai, Shanley Chong, Janice Tang, Bin Jalaludin, Margaret Thomas, Michelle Camilleri, Mandy Williams

## Abstract

**Background:** Childhood obesity is a major public health concern in Australia and the multicultural population of South-West Sydney. This study examined the influence of parents’ country of birth (COB) on the association between family socio-economic status (SES), assessed by area-level disadvantage and annual household income, and childhood obesity.

**Methods:** The analysis of data from the cross-sectional Growing Healthy Kids in Southwest Sydney (GHK-SWS) baseline survey of 1,815 children aged 5-16 years living in South Western Sydney, Australia employed generalised linear multinomial mixed models, with results presented as odds ratio and 95% confidence intervals.

**Results:** This study found a significant relationship between SES measures and the risk of overweight and obesity in the study population. The risks of childhood obesity are greatest in lower socioeconomic groups (across both SES metrics) regardless of their parents’ COB status. However, the overweight results are inconsistent across SES metrics and COB status. The risk of being overweight was lower for Australian-born parents living in disadvantaged areas but higher for those reporting lower income; and higher for Overseas-born parents living in disadvantaged areas but lower for those reporting low household income.

**Conclusion:** Parents’ SES is a consistent predictor of childhood obesity, and this relationship is not modified by parents’ COB. However, the likelihood of a child being overweight varied between the parents’ SES measures and COB. Prioritisation of the target population for area-level preventive public health interventions to modify obesogenic factors is recommended for a) obese children living in most disadvantaged areas and moderate disadvantaged areas regardless of their parents’ COB; b) overweight and obese children of moderate disadvantaged areas regardless of their parents’ COB.

## Introduction

Childhood obesity is a global public health issue, with significant health, economic and social effects. The global prevalence of childhood obesity is high, with substantial adverse immediate and long-term consequences^1^. A quarter of Australian children are overweight or obese, with no evidence of a decline in the prevalence^2^.

The aetiology of childhood overweight and obesity is multifaceted, involving a complex interaction between genetic, environmental, cultural, behavioural, and socio-economic factors^3^. Globally, low socio-economic status (SES) is an important determinant of the prevalence of childhood obesity^4^. The persistent relationship between low SES and increased overweight and obesity prevalence has been reported in several studies focusing on Australian youth^5,6^ as well as in children of other high-income countries^7^. Population data in Australia shows that differences in SES are related to the individual’s country of origin^8^, which is of particular significance in Australia, where approximately a third of the population is overseas-born or has overseas-born parents^9^.

There has been some progress in the investigation of obesogenic factors for children and other vulnerable populations, but studies are few and the results are inconsistent^10^. Parents’ country of birth (COB) and migration status^11,12^, mothers’ perceptions of healthy body weight^13^, maternal body mass index (BMI)^12^, and a range of SES indicators, including parents’ educational level, household income, place of residence, lifestyle patterns and employment status^14,15^ are important contributing factors to childhood overweight and obesity.

While previous research^8,16^ has found that both SES and ethnicity are associated with childhood obesity in Australian primary school children, less clear is the extent to which parents’ COB modifies the relationship between SES (measured by both area-level disadvantage and individual household income) and childhood overweight or obesity.

The high rates of childhood overweight and obesity in Australia—with one-third of children living in the South Western Sydney Local Health District (SWSLHD) region of New South Wales (NSW)^17^—has profound implications for public health policy, specifically concerning which groups of children should be prioritised for health interventions. The SWSLHD region has a population of approximately one million, including the largest population of culturally and linguistically diverse (CALD) communities in NSW, with 36% of residents born overseas and about 49% of families speaking a language other than English at home^18^. Such demographic characteristics make this region an ideal location for our study.

The objectives of this paper are twofold: a) to investigate the relationship between parents’ SES and childhood overweight and obesity in SWSLHD, and b) to determine whether this relationship is modified by parents’ COB.

## Methodology

### Ethics Approval & Formal Consent

This study was approved by the Human Research Ethics Committee of South Western Sydney Local Health District (HE18/078). Informed verbal consent was obtained from all the study participants (i.e., Parents or carers aged ≥18 years who could provide the maximum information about the selected child) before the interviews. The data was collected from October and November 2018. The data was accessed for research purposes from February 2021 and the authors had no access to information that could identify participants during or after data collection.

### Questionnaire and Study Sample

We analysed the baseline data from the GHK-SWS study to evaluate the study objectives. The GHK-SWS baseline study—a cross-sectional population-based survey of parents or carers of children aged 5-16 years in southwestern Sydney, NSW, Australia—was approved by the Southwest Sydney Local Health District Human Research Ethics Committee (HE18/078) and conducted in 2018, with a follow-up study planned to be conducted in 2027.

The main paper from the GHK-SWS study presents a detailed description of the study design and methodology^19^. In brief, the baseline questionnaire was developed using validated questions from the 2018 NSW Child Population Health Survey^20^, the NSW Schools Physical Activity and Nutrition Survey^21^ and the Make Healthy Normal campaign evaluation^22^. Survey domains included: demographics, health status, parental perception of height and weight, dietary consumption (including fruit and vegetables and discretionary foods), nutrition-related behaviours, physical activity participation and sedentary activity, as well as several knowledge and attitudes questions. The questionnaire was translated into Arabic, Vietnamese, and Cantonese/Mandarin (simplified Chinese) as these are the major language groups in our study area. GHK-SWS study data was collected using a computer-assisted telephone interview (CATI) system.

The target population included all households with children aged 5-16 years within the study area and the sample frame consisted of residents’ landline and mobile telephone numbers. The primary carer (mother or father or grandparents) who could provide maximum information about the selected child was interviewed from each randomly selected household. Following the contact, interviewers collected information on the number of eligible children living in the household and their gender and ages. If the household had more than one eligible child, then the interviewer randomly selected one of the eligible children using the CATI system’s random number generator.

## Measures

### Outcome measure

The main outcome measure was the child’s BMI. BMI was categorised using the age- and sex-specific International Obesity Task Force (IOTF) definitions^23^: thin/underweight (BMI<18 kg/m^2^), healthy weight (18 kg/m^2^ ≤BMI<25 kg/m^2^), overweight (25≤BMI<30 kg/m^2^) and obese (BMI≥30 kg/m^2^). To determine BMI, the respondent was asked two questions: 1. How tall is <CHILDNAME> without shoes? (in centimetres). 2. How much does <CHILDNAME> weigh without clothes or shoes? (in kilograms). As the aim of this paper was on childhood overweight and obesity, children with BMI <18 kg/m^2^ were excluded from the analysis (n=224).

### Predictors of interest

The study’s main predictor of interest was SES, which was assessed by two measures: area-level SES and household income of participants. Area-level SES was calculated from the 2016 Index of Relative Socio-Economic Disadvantage (IRSED)^24^ at the postcode level. The IRSED was created by the Australian Bureau of Statistics to compare social and economic disadvantage across geographic areas in Australia. The index is derived from Census variables such as income, educational attainment, unemployment, and people working in unskilled occupations. The IRSED was categorised into tertiles (Tertile 1: most disadvantaged/low SES; Tertile 2: middle disadvantaged/middle SES; and Tertile 3: least disadvantaged/high SES). The annual household income (before tax) of the respondents was grouped into four categories: $0-$51,999, $52,000-$77,999, $78,000-$103,999 and $104,000 or more.

### Effect Modifier

The other variable of interest was the parent’s COB (as an effect modifier), which was grouped into Australian-born and overseas-born. To determine COB, the respondent was asked – “Can you tell me in which country <CHILDNAME>s mother/father/you was/were born”? As we only focus on parents (mothers and fathers) as the main carer for the children, other care relationships (e.g., grandparents or relatives) were excluded from the analysis (n=51). In this paper, parent refers to mothers (73.5%) and fathers (21.5%) and only their responses to survey questions were included in the analysis.

Socio-demographic variables reported in the survey included both the child’s (age and gender) and parent’s demographic characteristics (educational attainment, annual household income, and rurality of residence using the Accessibility/Remoteness Index of Australia Plus (ARIA+) index. The rurality of residence quantifies remoteness in terms of the travelling distance to different-sized population-adjusted service centres^25^. The ARIA+ scores are classified into five groups: major cities, inner regional, outer regional, remote, and very remote.

Additionally, due to the known relationship between healthy lifestyle behaviours, childhood BMI and area-level SES^5,26^, three health behaviour factors were explored as potential mediators: physical activity (PA) levels, sedentary screen time behaviour (SB) and consumption of sugar-sweetened beverages (SSB). For PA, children were categorised according to whether they met the current Australian PA guidelines^27^ (yes, no) and were asked the following question: “Over the past 7 days, on how many days was <CHILDNAME> engaged in moderate to vigorous physical activity for at least 60 minutes each day (this can be accumulated over the entire day, for example in bouts of 10 minutes)?” For SB, the current Australian guidelines recommend that children aged 5-17 years limit sedentary recreational screen time (such as watching TV, iPad, DVDs or on a mobile phone) to two hours per day^27^. SB was assessed by asking participants: “In a typical week (Monday to Sunday), how many hours in total does <CHILDNAME> spend watching TV/iPad/DVDs, using the computer/iPad/mobile phone for fun or playing video games?” and categorised into <2 hours of screening time per day and >2 hours of screening per day. SSB consumption was assessed by asking participants” How many cups of soft drink/cordial/sport drink/energy drink, does their child usually drink in a day? Total consumption of SSB per day was categorised into no cups, ≤1 cup, and >1 cup per day. The current Australian Dietary Guidelines discourage the consumption of any SSBs in children due to the numerous detrimental health effects^28^.

### Data analysis

A directed acyclic graph (DAG) was used to identify potential confounders, mediators, and effect modifiers in the relationship between SES and children’s overweight and obesity (Figure 1). Following a comprehensive descriptive analysis, the R package “mma” was used to conduct multiple mediation analysis^29^. Multiple mediation analysis examined the presence of the potential mediator factors: meeting PA, SB and SSB consumption guidelines. The algorithms for the estimation of indirect effects were repeated 50 times with 500 times of bootstrap resampling. These three health behaviours were not significant mediators for children’s overweight and obesity and were included as confounders in the final model. Proc GLIMMIX procedure in SAS v9.4 was then used to conduct generalised linear multinomial mixed models^29^. This included random intercepts, which accounted for Local Government Area (LGA) clustering effects, to determine associations between area-level SES and Children’s overweight and obesity. Children’s age, gender, parent’s relationship to child (as mother or father), parent’s educational attainment, annual household income, and rurality of residence were included as confounders in the final model. Effect modification by the main carer’s (mother or father) COB was tested and was statistically significant (p<0.0001).

**Figure 1:**
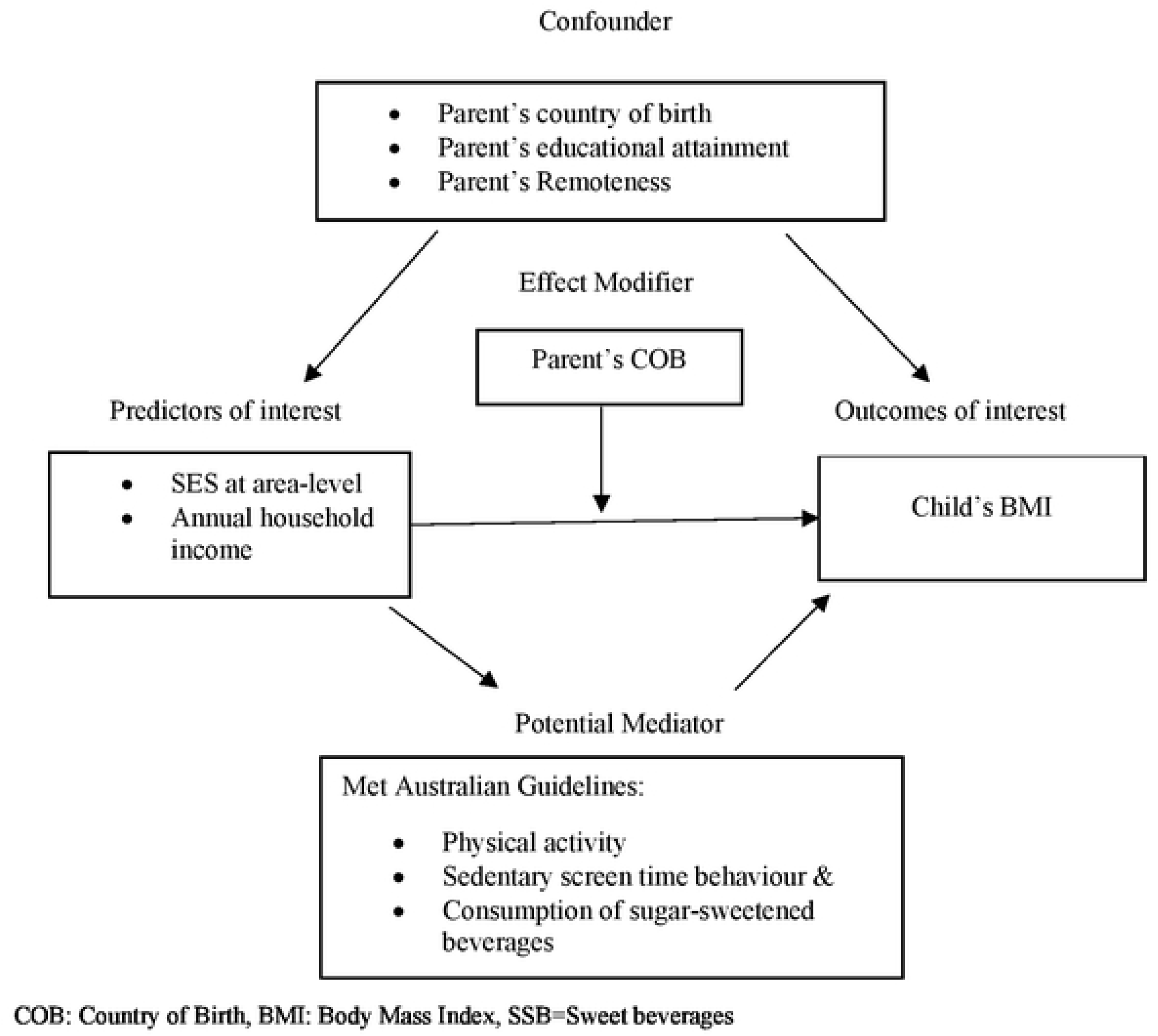
Directed acyclic graph of the relationship between socioeconomic status and body mass index.

Results from the multinomial multilevel regression model are presented as odds ratio (ORs) and associated 95% confidence intervals (95%CIs). Survey weighting was used in all analyses to ensure the survey sample was representative of all children living in SWSLHD.

## Results

The socio-demographic characteristics of participants are presented in Table 1. In total, the GHK-SWS study collected data from 1,815 children. After exclusion of thin (underweight) children (n=224) and other non-parents (n=51), the study findings are from the analysis of 1,535 children. Overall, 57% of children were within the healthy weight range (n=878), 23% were overweight (n=367), and 20% were obese (n=290). About half of the participants were boys (51%, n=790) and the mean age was 10.8 years (SD=4.5 years). The majority lived in major cities (92%), with more than half of the participants (57%) living in more disadvantaged areas and about one-third (34%) reporting an annual household income of less than $52,000. Most participating parents were mothers (74%), about half of whom were born in Australia (51%), 35% have completed a university degree, 32% with a Technical and Further Education (TAFE) diploma, and 33% with ≤12 years of school education.

**Table 1:** Participants sociodemographic characteristics, n=1535.

| Variable | Level | n, % |
| --- | --- | --- |
| **Age** | 5-11 years | 890, 58.0 |
|  | 12-16 years | 645, 42.0 |
| **Gender** | Boys | 790, 51.2 |
|  | Girls | 745, 48.8 |
| **Parent's relationship** | Mother | 1096, 73.5 |
|  | Father | 439, 26.5 |
| **Parent's country of birth (missing=45)** | Australia | 780, 50.7 |
|  | Other | 755, 49.3 |
| **Parent's educational attainment**<br>**(missing=7)** | <=Year 12 | 430, 33.1 |
|  | Trade, TAFE, or Diploma | 523, 32.0 |
|  | University degree | 575, 34.9 |
| **Annual household Income (before tax)**<br>**(missing=232)** | \$0-\$51,999 | 354, 34.2 |
| | \$52,000-\$77,999 | 254, 20.5 |
| | \$78,000-\$103,999 | 250, 16.3 |
| | \$104,000 or more | 445, 28.9 |
| **Location** | Major Cities | 1404, 91.7 |
|  | Inner Regional | 131, 8.3 |
| **Area-Level socio-economic status** | Most disadvantaged | 843, 57.0 |
|  | Moderate disadvantaged | 263, 16.6 |
|  | Least disadvantaged | 429, 26.4 |
| **Body mass index** | Healthy weight | 878, 57.2 |
|  | Overweight | 367, 22.5 |
|  | Obese | 290, 20.3 |
| **Met physical activity guideline** | No | 1212, 77.7 |
|  | Yes | 323, 22.3 |
| **Met sedentary behaviour guideline (missing=17)** | No | 349, 22.2 |
|  | Yes | 1169, 77.8 |
| **Sugar sweetened beverages (per day)** | No cups | 786, 52.4 |
|  | 1 cup | 510, 32.4 |
|  | 2+ cups | 239, 15.2 |

TAFE: Technical and Further Education

### Socio-demographic factors and BMI

In an adjusted model, using healthy weight as the reference, the socio-demographic factors associated with higher odds of being overweight in children were: girls, aged 5-11 years, parents born in Australia, parents who had only completed year 12 or less of education, families with a reported annual household income less than $77,999, living in a major city and a moderate disadvantaged area (Table 2). Children were more likely to be obese if they were: boys, aged 5-11 years, their fathers were the main carers, parents were born overseas, parents who had not completed a university degree, from families with a reported annual household income of less than $52,000, living in inner regional areas, and from the most disadvantaged areas. (Table 2).

**Table 2:** Adjusted associations between socio-demographic factors, health behaviours and overweight and obesity*.

| Variable | Level | Healthy weight<br>n, % | Overweight<br>n, % | Adjusted OR, 95% CI | Obese<br>n, % | Adjusted OR, 95% CI |
| --- | --- | --- | --- | --- | --- | --- |
| **Age** | 5-11 years | 483,<br>53.0 | 191,<br>20.6 | 1 | 216,<br>26.4 | 1 |
|  | 12-16 years | 395,<br>63.0 | 176,<br>25.1 | 0.83,<br>0.81-0.86 | 74,<br>11.9 | 0.28,<br>0.27-0.29 |
| **Gender** | Boys | 435,<br>55.6 | 201,<br>22.4 | 0.98,<br>0.95-1 | 154,<br>22.0 | 1.04,<br>1.01-1.07 |
|  | Girls | 443,<br>58.9 | 166,<br>22.6 | 1 | 136,<br>18.5 | 1 |
| **Parent's relationship** | Mothers | 643,<br>47.8 | 254,<br>22.4 | 1 | 199,<br>19.9 | 1 |
|  | Fathers | 235,<br>55.6 | 113,<br>22.8 | 0.99,<br>0.96-1.02 | 91,<br>21.5 | 1.18,<br>1.14-1.22 |
| **Parent's country of birth** | Australia | 466,<br>60.1 | 185,<br>22.5 | 1 | 129,<br>17.4 | 1 |
|  | Other | 412,<br>54.2 | 182,<br>22.4 | 0.88,<br>0.85-0.91 | 161,<br>23.3 | 1.43,<br>1.38-1.48 |
| **Parent's educational attainment** | <=Year 12 | 219,<br>51.3 | 112,<br>23.4 | 1.1,<br>1.06-1.14 | 99,<br>25.3 | 1.99,<br>1.91-2.08 |
|  | Trade, TAFE, or Diploma | 305,<br>57.8 | 116,<br>21.5 | 1.02,<br>0.99-1.06 | 102,<br>21.0 | 1.49,<br>1.43-1.55 |
|  | University degree | 351,<br>62.9 | 138,<br>22.4 | 1 | 86,<br>14.8 | 1 |
| **Annual household Income** | \$0-\$51,999 | 175,<br>46.6 | 86,<br>22.8 | 1.3,<br>1.24-1.35 | 93,<br>30.6 | 1.66,<br>1.59-1.74 |
| | \$52,000-\$77,999 | 146,<br>60.0 | 58,<br>22.9 | 1.09,<br>1.05-1.14 | 50,<br>17.2 | 0.92,<br>0.88-0.97 |
| | \$78,000-\$103,999 | 145,<br>61.4 | 62,<br>22.4 | 1,<br>0.95-1.05 | 43,<br>16.2 | 0.93,<br>0.88-0.98 |
| | \$104,000 or more | 271,<br>62.9 | 108,<br>21.3 | 1 | 66,<br>15.8 | 1 |
| **Location** | Major Cities | 800,<br>56.5 | 338,<br>22.9 | 1 | 266,<br>20.6 | 1 |
|  | Inner Regional | 78,<br>65.1 | 29,<br>17.6 | 0.68,<br>0.62-0.75 | 24,<br>17.3 | 3.11,<br>2.83-3.43 |
| **Area-level SES** | Most disadvantaged | 454,<br>53.6 | 209,<br>22.6 | 0.95, 0.91-0.99 | 180,<br>23.8 | 1.49, 1.42-1.57 |
|  | Middle disadvantaged | 147,<br>56.5 | 67,<br>23.8 | 1.27,<br>1.21-1.34 | 49,<br>19.6 | 1.74,<br>1.63-1.84 |
|  | Least disadvantaged | 277,<br>65.4 | 91,<br>21.5 | 1 | 61,<br>13.1 | 1 |
| **Met physical activity guideline** | No | 686,<br>56.4 | 301,<br>24.3 | 1.42,<br>1.36-1.47 | 225,<br>19.4 | 0.92,<br>0.89-0.96 |
|  | Yes | 192,<br>60.1 | 66,<br>16.4 | 1 | 65,<br>23.5 | 1 |
| **Met sedentary behaviour guideline** | No | 181,<br>51.2 | 95,<br>24.5 | 1.13,<br>1.09-1.17 | 73,<br>24.4 | 1.49,<br>1.44-1.55 |
|  | Yes | 686,<br>58.7 | 270,<br>22.2 | 1 | 213,<br>19.1 | 1 |
| **Sugar sweetened beverages (per day)** | No cups | 481,<br>60.3 | 171,<br>21.6 | 1 | 134,<br>18.1 | 1 |
|  | >0-<1 cup | 287,<br>58.6 | 124,<br>22.2 | 1.22,<br>1.18-1.26 | 99,<br>19.2 | 1.37,<br>1.32-1.42 |
|  | 1+ cups | 110,<br>43.5 | 72,<br>26.2 | 1.52,<br>1.45-1.59 | 57,<br>30.3 | 2.52,<br>2.42-2.63 |

\*Type 3, p-value for each variable is <.0001
SES= Socio Economic Status

### Health Behaviours and BMI

In the same adjusted model (Table 2), children are more likely to be overweight when PA and SSB guidelines are not met. Similar patterns were observed for children with higher odds of being obese except for meeting PA guidelines: children who meet PA guidelines are more likely to be obese (OR=1.09, 95 % CI=1.04-1.12) compared to all other children.

### SES and BMI by Parents’ COB

Figure 2 shows the associations between area-level SES, annual gross household income and child overweight and obesity by parents’ COB after adjusting for confounders.

**Figure 2:**
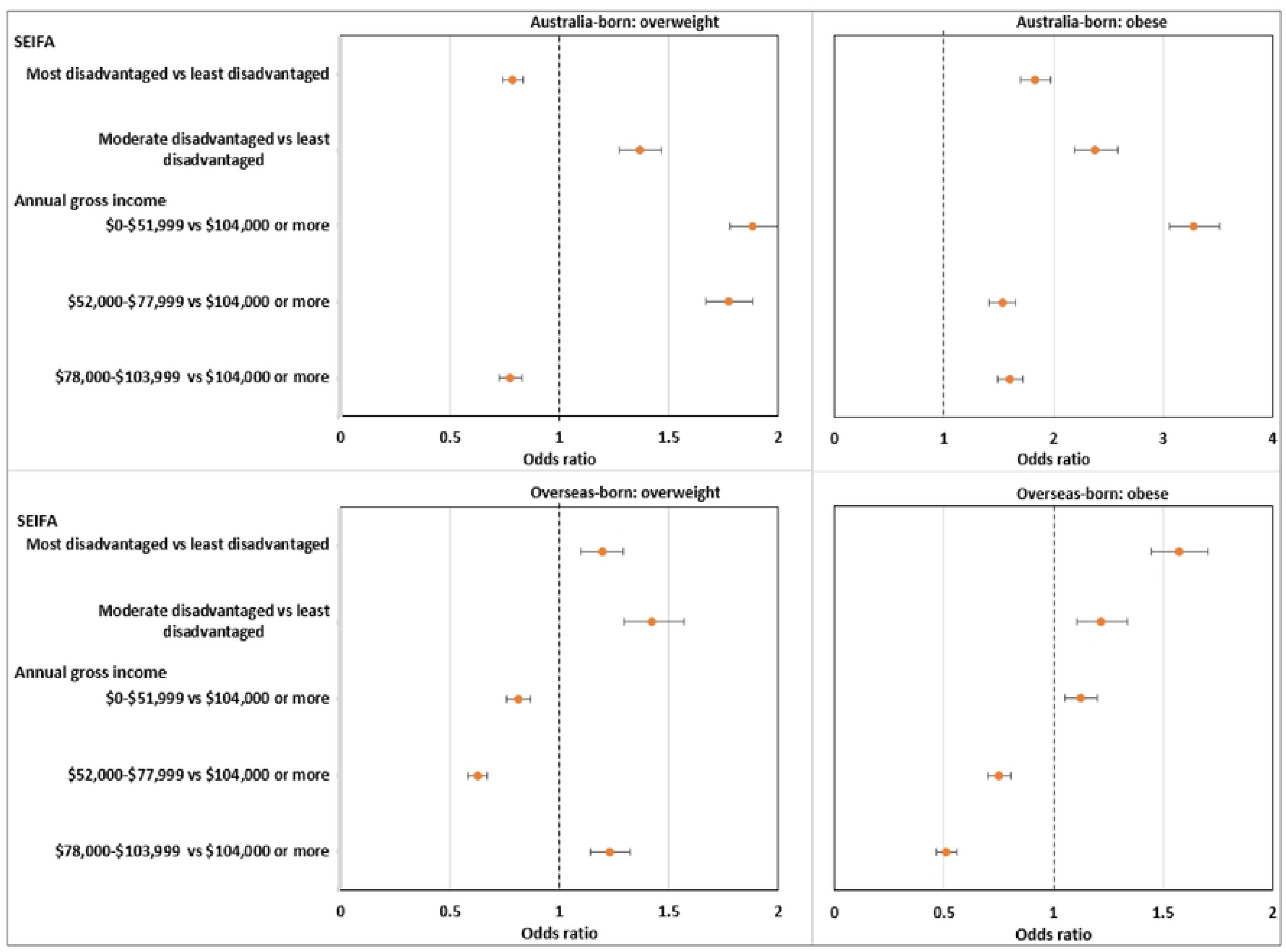
Association between IRSD, annual gross household income and BMJ by parent’s country of birth after adjusting for confounders1, adjusted OR 95% CI. ^1^Age, sex, parent’s relationship with child (mother or father) and educational attainment, geographical location, sugar sweetened beverages consumption, whether met physical activity guideline, whether met sedentary behaviour guideline were included as covarietes

### Overweight and obesity in children with parents born in Australia

If parents were born in Australia, the odds of their child being overweight were highest for families living in moderate disadvantaged areas (OR=1.36, 95% CI=1.27-1.46) but lowest for families living in the most disadvantaged areas (OR=0.78, 95% CI=0.74-0.83), compared to those living in the least disadvantaged areas. Compared to an annual household income of $104,000 or more, an annual household income below $77,999 was associated with a higher risk for overweight ($0-$51,999: OR=1.88, 95% CI=1.78-2.00; $52,000-$77,999: OR=1.77, 95% CI=1.67-1.88), while an annual household income of $78,000-$103,999 was associated with a lower risk (OR=0.78, 95% CI=0.73-0.83).

The risk of a child being obese was higher among participants living in the most and moderately disadvantaged areas (most disadvantaged areas: OR=1.83, 95% CI=1.70-1.97; moderate disadvantaged areas: OR=2.38, 95% CI=2.19-2.58) than living in the least disadvantaged areas. The odds of being obese were also higher for all three annual household income categories compared to the reference category of >$104,000: ($0-$51,999: OR=3.28, 95% CI=3.06-3.52; $52,000-$77,999: OR=1.53, 95% CI=1.41-1.66; $78,000-$103,999 OR=1.60, 95% CI=1.49-1.72).

### Overweight and obesity in children with parents born overseas

About parents born overseas, the odds of their children being overweight were higher for families living in the most disadvantaged areas (OR=1.19, 95% CI=1.10-1.29), and moderate disadvantaged areas (OR=1.42, 95% CI=1.29-1.57). The odds of children being overweight were lower among families reporting an annual household income below $77,999 than those above $104,000 or more ($0-$51,999: OR=0.81, 95% CI=0.76-0.87; $52,000-$77,999: OR=0.62, 95% CI=0.58-0.67). However, the odds of children being overweight were higher among families with a household annual income of $78,000-$103,999 (OR=1.23, 95% CI=1.14-1.32).

As area-level SES increased, the odds of these children being obese followed a clearly increasing trend from least disadvantaged to most disadvantaged (moderate disadvantaged areas: OR=1.22, 95% CI=1.11-1.34; most disadvantaged areas: OR=1.57, 95% CI=1.45-1.70).).

The odds of children being obese were higher among families with a reported annual household income of $0-$51,999 (OR=1.12, 95% CI=1.05-1.20), though to a much lesser extent than children whose parents were born in Australia. However, the odds of children being obese were lower among participants whose annual household income was between $52,000 and $103,999 ($52,000-$77,999: OR=0.75, 95% CI=0.70-0.80; $78,000-$103,999: OR=0.51, 95% CI=0.47-0.56).

## Discussion

This study found a significant relationship between SES measures (assessed by area-level disadvantage and annual household income) and the risk of overweight and obesity among children aged 5-16 years living in Southwest Sydney. The obesity results are consistent across SES metrics and COB groups. A higher risk for childhood obesity was found for children in low SES households regardless of whether SES status was measured by household income or area of residence and this relationship held whether the responding parent was born in Australia or outside Australia. Childhood obesity risk was three-fold higher for Australian-born parents with the lowest household income level.

The overweight results, however, are inconsistent across SES metrics and COB groups. Children of Australian-born parents had a lower risk of being overweight if they lived in low-SES areas, but children of overseas-born parents had a higher risk of being overweight if they lived in low-SES areas. When SES was measured by annual household income, the opposite was found. Children of Australian-born parents with low annual household incomes had a higher risk of being overweight whereas children of overseas-born parents with low annual household incomes had a lower risk of being overweight.

The increased risk of obesity in children of low SES groups across both Australian and overseas-born parents in our study accords with the recent World Health Organization (WHO) report on ending childhood obesity^30^. The existing evidence from WHO and recent evidence from Australia demonstrates that the risks of childhood obesity are highest in lower SES groups living in high-income countries^31,32^. Indeed, in high-income countries, children from low SES population groups such as immigrants, indigenous, and culturally and linguistically diverse groups, are at a particularly high risk of developing obesity^30^.

While many studies have examined the contribution of SES to differences in obesity by ethnic group, very few studies have considered migration status as a factor^33^. The limited evidence from studies conducted in Europe^34^, USA^35^, Australia^36^ and Germany^37^ suggests that weight increases substantially among migrant children over a post-migration period of 10 to 15 years and social and economic disadvantage of migrants is linked to an increased risk of childhood obesity, an association our study findings confirm.

In this study, the influence of SES as measured by both area and household income metrics on childhood overweight and obesity was inconsistent across the two SES measures. SES measured by household income and area-level disadvantage provide different contextual information. Further, the risk of childhood overweight and SES is also influenced by the type of SES measure used in the analysis. For example, children of immigrants with low SES were at higher risk of being overweight when parental education was used as a proxy for SES^37^ but not when employment was used as an indicator of SES^38^. Area-level SES also reflects other area-level characteristics such as access to healthy foods and facilities for physical activity, employment opportunities, and crime levels. Therefore, while household income is an important SES metric, area-level SES may be a more useful measure when policy decisions must be made to allocate resources or to implement population-level programs. The results from our study and the literature indicate the need for further detailed studies to understand the complexity and variation of childhood overweight and obesity in subgroups of the population.

While household income is a known predictor of childhood obesity, this association was significantly modified by parents’ country of birth. A systematic review of socioeconomic inequalities in childhood obesity in the United Kingdom, for example, found that while household income can reliably be ascribed as a determinant of childhood obesity, such findings were not consistent across all studies^39^. Our study also confirms that low household income is a reliable predictor of childhood obesity, regardless of children’s COB status. Still, when it comes to the question of being overweight, parents’ COB is a modifying factor. Future studies would benefit from the inclusion of both area- and individual-level SES metrics to explore their value in predicting childhood overweight and obesity.

Childhood overweight and obesity arise from exposure of the child to an obesogenic environment and insufficient responses to that environment. Such responses vary from person to person for reasons that may be social, cultural, economic, environmental, or behavioural, as discussed below.

There are plausible explanations for the increased risk of childhood overweight and/or obesity in the children of overseas-born parents living in disadvantaged neighbourhoods. Residents of disadvantaged areas are more likely to be living with low wages, unemployment or, uncertain employment, particularly if they are recent migrants, females, single parents with dependent children, or speak a language other than English at home^40,41^. Subsequent generations of migrants, even those with higher income levels, are also more likely to experience marginalisation and reduced opportunity^42^, attain lower educational levels, and lack the resources to own property^43^. Moreover, while migrants may arrive in their new country with certain health advantages, they are more likely to experience social isolation or marginalisation, stress, and weight gain 10-15 years after migration^33^.

Besides the socioeconomic disadvantage of migrants, their exposure to obesogenic environmental factors in host countries may also influence the higher prevalence of overweight/obesity among their children^44,45^. This phenomenon has been linked to the challenges of acculturation, including lifestyle changes in the host country, lower levels of physical activity, increased sedentary lifestyle^18,34^ and the adoption of unhealthy diets^45–47^.

## Strengths and Limitations

BMI based on self-reported height and weight may cause moderate measurement bias. However, evidence from a previous study suggests that correlations between self-reported height, weight and BMI values and objectively measured values remained high and hence support the use of self-reported values^48^. While the study provides comprehensive information based on sufficient sample size, representative population data and a standardised questionnaire, a potential limitation is the assessment of the causal association between SES, parents’ country of birth and childhood overweight and obesity, because of its cross-sectional design. While children’s health behaviours, such as meeting physical activity, sedentary behaviour, and sugar-sweetened beverage consumption guidelines, were adjusted in the final model, future studies that include data on other obesogenic factors (such as the availability of unhealthy food stores) would be an important area of research.

## Conclusion

Parents’ SES (measured by area level and household income) is a consistent predictor of childhood obesity, and this relationship is not modified by parents’ COB. However, the likelihood of being overweight varied between the SES measures and parents’ COB. It is recommended that the priority populations that should be targeted for preventive public health interventions to modify obesogenic factors include a) obese children living in most disadvantaged regardless of their parents’ COB; b) overweight and obese children of moderate disadvantaged areas regardless of their parents’ COB.

Identifying the causes underlying the prevalence of overweight in children from different COB backgrounds can provide further insight into the development of effective and relevant policies and health promotion initiatives. As both area-level and household-level SES metrics are associated with childhood obesity but inconsistently, therefore, future studies should include both metrics when exploring associations with childhood obesity.

## Data Availability

Data available through manuscript.

## Acknowledgements

We firstly acknowledge the parents and carers of children living in the South West Sydney Local Health District who generously gave their time to complete the GHK-SWS Survey. We acknowledge the contribution of McNair Yellow Squares who were contracted by the University of Sydney to undertake the survey. We also acknowledge the Growing Healthy Kids Evaluation Advisory Group which contributed to the development of the survey and analysis of the data. Dr Phil Hughes of Engine Group provided the data weighting. We would like to also thank Francis Fox for his language edits.

## Authors’ contributions

VK wrote the manuscript and was responsible for the design, drafting, interpretation of data, reviewing and revisions of the manuscript. SC contributed to the design, conducted analysis of data, wrote the results section, reviewing and revisions of the manuscript. BJ contributed to the design, methodology, results and extensive reviewing, and revisions of the manuscript and oversaw the data analysis and interpretation. JT, MC, MT, and MW contributed to the revisions of the manuscript. All authors read and approved the final manuscript.

